# Device-measured moderate-to-vigorous physical activity and prognosis of inflammatory bowel disease

**DOI:** 10.1101/2024.10.30.24316417

**Authors:** Zixuan He, Yuhao Sun, Hanyi Huang, Yilong Liu, Lintao Dan, Xixian Ruan, Tian Fu, Zhaoshen Li, Xiaoyan Wang, Jie Chen, Yu Bai

## Abstract

**Importance:** Evidence on physical activity recommendations for inflammatory bowel disease (IBD) patients is limited, with concerns about high-intensity activity.

**Objective:** To investigate the associations of accelerometer-measured moderate-to-vigorous physical activity (MVPA) with bowel resection risk and mortality among individuals with IBD.

**Design, setting, and participants:** This longitudinal cohort study collected data from 1,303 UK Biobank participants with IBD diagnosis and device-measured physical activity from 2013 to 2015, with follow-up data examined until 2022.

**Exposures:** MVPA was classified based on data measured by wrist-worn accelerometers over a 7-day period. MVPA patterns were defined as inactive (< 150 minutes/week), active weekend warrior (≥ 150 minutes/week, ≥50% of total MVPA achieved in 1-2 days), and regularly active (≥ 150 minutes/week, not active weekend warrior).

**Main Outcomes and Measures:** The bowel resection was identified through operational records from hospital inpatient data of the National Health Service. Deaths were documented through linkage to the national death register. Associations were estimated using multivariable Cox regression models and nonlinearity was assessed by restricted cubic spline.

**Results:** During a median follow-up of 7.8 years, 56 incident bowel resection cases and 86 deaths were documented. After multivariable adjustment, participants in the highest tertile of MVPA duration had lower risks of bowel resection (HR, 0.44; 95% CI, 0.22-0.86) and mortality (HR, 0.49; 95% CI, 0.27-0.89) compared to those in the lowest tertile. MVPA duration is linearly associated with bowel resection (P _non-linear_ = 0.13) while its dose-response relationship with mortality plateaus at approximately 58 min/day (P _non-linear_ = 0.02). Regarding MVPA patterns, the active weekend warrior pattern was inversely associated with bowel resection risk (HR, 0.28; 95% CI, 0.12-0.65), the regularly active pattern was inversely associated with both bowel resection risk (HR, 0.37; 95% CI, 0.19-0.69) and mortality (HR, 0.53; 95% CI, 0.31-0.91) compared to the inactive.

**Conclusion and relevance:** Longer accelerometer-measured MVPA was associated with reduced bowel resection risk and mortality. The regularly active pattern may be the optimal choice for individuals with IBD, while the active weekend warrior pattern still provides health benefits compared to being inactive.

## Introduction

Inflammatory bowel disease (IBD), comprising Crohn’s disease (CD) and ulcerative colitis (UC), is a chronic inflammatory condition of the gastrointestinal tract that significantly impairs patients’ quality of life and is associated with considerable morbidity.^1^ Managing IBD typically requires long-term medication, with many patients eventually requiring surgical intervention.^2^ However, these medical treatments are not effective for all patients, and they impose a significant economic burden on both individuals and healthcare systems.^3,4^ A previous study reported that 30%–50% of patients with IBD seek complementary and alternative medicines including physical activity.^5^ Recognized for its ability to reduce systemic inflammation and promote metabolic health, physical activity is a crucial modifiable lifestyle factor recommended for a range of chronic non-communicable diseases.^6,7^ The World Health Organization recommends at least 150 minutes of moderate-to-vigorous physical activity (MVPA) per week to achieve overall health benefits.^8^ However, the applicability of this guideline to individuals with IBD remains uncertain. Some recommendations for the general population may not be directly relevant to IBD patients; for example, previous research indicated that smoking cessation, typically beneficial for health, is associated with worsening disease activity in patients with ulcerative colitis.^9^ Moreover, the specific symptoms associated with IBD and related psychological factors may significantly impact patients’ activity patterns and limit their participation.^10–12^

Current literature indicates a link between exercise and IBD, suggesting that regular physical activity may benefit patients by releasing protective myokines, inducing an anti-inflammatory environment, improving gut microbiota, and reducing the risk of IBD-specific complications such as osteoporosis.^13,14^ However, concerns exist regarding excessive and strenuous physical activity, which might exacerbate gastrointestinal symptoms.^5^ Existing guidelines lack consensus on specific exercise recommendations regarding intensity, duration, and pattern. Only one guideline encourages clinicians to assess patients’ activity levels, identify and address barriers to physical activity, and promote increased activity within patients’ tolerance limits.^15^ The paucity of studies on the role of physical activity among IBD patients poses challenges for healthcare providers and patients in determining optimal exercise regimens for managing the disease. To be specific, current studies primarily focus on short-term outcomes, offering limited insight into long-term prognostic outcomes such as surgery and mortality. Furthermore, no high-quality evidence exists confirming whether differences exist between exercise patterns. In addition, previous studies often had limited sample sizes and primarily relied on self-reported data to evaluate physical activity, which is susceptible to recall bias and may not accurately reflect actual activity levels. ^13,16,17^

To provide a more comprehensive understanding of the role of MVPA on IBD prognosis and overcome the limitations of self-reported physical activity, this study investigated whether higher duration and specific patterns of objectively measured MVPA are associated with a reduced risk of bowel resection surgery and mortality in individuals with IBD.

## Methods

### 1.1 Study population

Participants in this study were from the UK Biobank, a large prospective cohort designed to allow detailed investigation of risk factors for a wide range of diseases. It involved half a million participants aged 40-69 years at recruitment in 2006-2010.^18,19^ Participants were initially assessed at 22 assessment centers across the UK via touchscreen questionnaires, face-to-face interviews, physical measurements, and biological samples. Additional assessments were conducted to enhance phenotyping, including web-based questionnaires and accelerometry. Embedded within the UK’s National Health Service (NHS), the UK Biobank tracks the long-term health of participants and identifies disease outcomes using routine medical records.^20^ The study of the UK Biobank received general ethical approval from the National Health Service National Research Ethics Service (Ref 21/NW/0157). All participants provided written informed consent.

Our study initially recruited 103,648 participants with accelerometer data. After excluding 9,169 participants with insufficient accelerometer data quality and 93,176 participants without baseline IBD, a total of 1,303 individuals with both valid device-measured physical activity and baseline IBD were included (**Figure S1**). This study followed the STROBE guidelines for cohort studies.

### 1.2 Measurement of MVPA

Physical activity was measured using wrist-worn Axivity AX3 triaxial accelerometers, deployed between February 2013 and December 2015. Participants were randomly invited via email to wear an Axivity AX3 wrist-worn triaxial accelerometer on their dominant wrist for 7 consecutive days.^21,22^ This device has been validated in a previous study using multi-axis shaking tests and showed equivalent output to GENEActive accelerometer^23^ which has been validated in both laboratory and free-living assessments.^24,25^ Triaxial acceleration data is collected at 100Hz with a dynamic range of ±8g and extracted after calibration, flagging and resampling, removing gravity and noise, and identifying non-wear periods.^22^ Further, accelerometer data was classified into movement behaviors (sleep, sedentary behavior, light physical activity, and MVPA) using machine-learning methods that have been validated in the UK sample.^26^ MVPA is defined as the physical activity performed at over 3 metabolic equivalents of task (METs), where 1 MET represents the energy expenditure of an individual at rest. Non-wear periods are imputed using time-matched averages from other days to account for potential diurnal bias in wear patterns and a physical activity outcome variable is finally constructed by averaging all worn and imputed values for each participant.^22^ According to prior research, 1-week accelerometer measurements have moderate to high correlations (intraclass correlation coefficients = 0.54-0.82) with repeated assessments over 6 months to 4 years.^27,28^

MVPA duration was categorized into tertiles: low, medium, and high. MVPA patterns were classified based on guideline-based threshold^8^ into inactive (<150 minutes/week), active weekend warrior (≥150 minutes/week with ≥50% of MVPA achieved in 1-2 days), and regularly active (≥150 minutes/week but not active weekend warrior).^29,30^

### 1.3 Outcome Assessment

The bowel resection was identified through operational records from hospital inpatient data of the National Health Service (NHS). All operations and procedures are recorded using OPCS-4 (Offices of Population, Censuses and Surveys: Classification of Interventions and Procedures version 4) codes. Corresponding OPCS-4 codes are provided in previous studies.^31,32^ Deaths were documented through linkage to the national death register. Follow-up began on the date when all accelerometer measurements were completed and continued until the first occurrence of bowel resection, death, loss to follow-up, or the last date of hospital admission (October 31, 2022, for England; August 31, 2022, for Scotland; and May 31, 2022, for Wales).

### 1.4 Assessment of Covariates

Covariates were selected based on prior knowledge,^31^ including age in years, sex (male/female), ethnicity (White/others), education attainment (college and above/below college), Townsend deprivation index (TDI), employment (employed/unemployed), body mass index (BMI), smoking status (ever/never), drinking status (current/non-current), adherence to a healthy diet (yes/no). TDI was derived from participants’ postcodes, with a higher score indicating greater deprivation. BMI was calculated as body mass divided by the square of the body height (kg/m^2^). Adherence to a healthy diet was defined as meeting at least 4 out of 7 criteria related to the consumption of fruits, vegetables, fish, unprocessed red meats, whole grains, refined grains, and processed meats. ^33,34^ In sensitivity analyses, additional covariates were accounted for, including C-reactive protein, self-reported overall health (good, fair, or poor), waist-hip ratio (WHR), and common IBD-related medications (use/not use) comprising 5-aminosalicylic acid, corticosteroids, and immunosuppressors.^31^ Detailed information on covariates is presented in Supplementary **Table S1**.

### 1.5 Statistical analysis

Baseline characteristics by tertile of MVPA duration in minutes per day are presented as means (SDs) for continuous variables and counts (percentages) for categorical variables. Missing values (<0.84% for all covariates) were imputed using the median for continuous variables and mode for categorical variables (**Table S1**). Associations between duration and patterns of MVPA and IBD prognosis were estimated using two multivariable Cox regression models. Model 1 was adjusted for age and sex. Model 2 was further adjusted for ethnicity, education, TDI, employment, BMI, smoking status, drinking status, and adherence to a healthy diet. The proportional hazards assumption was tested based on Schoenfeld residuals (all models satisfied the assumption with *P*>0.15). We used E-value to assess the potential impact of unmeasured confounding on observed associations from main models.^35^ The potential non-linear association between MPVA duration and IBD prognosis was tested using the restricted cubic spline with three knots placed at the 10th, 50th, and 90th percentiles of daily MVPA duration and 0 was used as a reference.^36^ Nonlinearity in exposure-outcome relationships was evaluated by likelihood ratio tests. Subgroup analyses assessed interactions by age (>60 years/≤60 years), sex (male/female), and time since IBD diagnosis (>15 years/≤15 years), with multiplicative and additive interactions evaluated through interaction terms and relative excess risk due to interaction (RERI), respectively.^37^ Additionally, we explored the associations of MVPA and IBD prognosis in subtypes of IBD (i.e. CD and UC).

Several sensitivity analyses were performed to further test the robustness of the results: (1) we employed multiple imputation, a distinct approach from the primary analysis’s imputation strategy, to address missing values; (2) participants who had the outcome of interest within one year of follow-up were excluded to minimize reverse causation; (3) separate analyses were performed with additional adjustments made for C-reactive protein, self-reported health, WHR, IBD-related medications, and baseline bowel resection history (for analyses of bowel resection), respectively. All analyses were performed using R software, version 4.2.1. A two-tailed *P* <0.05 was considered statistically significant.

## Results

During a median follow-up period of 7.8 years, we recorded 56 bowel resections and 86 all-cause deaths of the 1,303 individuals with IBD included. Baseline characteristics of participants stratified by tertile of MVPA duration and pattern are shown in **Table 1** and **Table S2**, respectively. 506 (38.8%) participants were classified as inactive, 287 (22.0%) as active weekend warriors, and 510 (39.1%) as regularly active. Individuals who undertook more MVPA were generally younger, more likely to be male, employed, had higher educational attainment, lower BMI, lower inflammation level, and better self-reported health. **Figure 1A** shows the distribution of MVPA duration across different MVPA patterns. The median MVPA time was 64.6 minutes/week for the inactive group, 380.8 minutes/week for active weekend warriors, and 271.3 minutes/week for regular exercisers.

**Table 1.** Baseline characteristics of participants stratified by tertile of moderate-to-vigorous physical activity duration.

| Characteristics | Overall (n=1303) | MVPA duration, min/day |  |  |
| --- | --- | --- | --- | --- |
|  |  | Tertile 1 (>0 to ≤17)<br>(n=435) | Tertile 2 (>17 to ≤40.7)<br>(n=435) | Tertile 3 (>40.7)<br>(n=433) |
| Age, year | 56.6 (7.7) | 57.3 (7.7) | 56.5 (8.0) | 55.9 (7.4) |
| Female (%) | 701 (53.8) | 277 (63.7) | 246 (56.6) | 178 (41.1) |
| White (%) | 1267 (97.3) | 424 (97.7) | 419 (96.3) | 424 (97.9) |
| College and above (%) | 478 (37.0) | 128 (29.6) | 152 (35.3) | 198 (46.0) |
| Townsend deprivation index | -1.60 (2.88) | -1.61 (2.92) | -1.51 (2.97) | -1.69 (2.74) |
| Employed (%) | 799 (61.5) | 246 (56.8) | 269 (61.8) | 284 (65.7) |
| Body mass index, kg/m<sup>2</sup> | 26.81 (4.66) | 28.44 (5.49) | 26.55 (4.24) | 25.45 (3.54) |
| Ever smoking (%) | 655 (50.3) | 247 (56.9) | 205 (47.1) | 203 (46.9) |
| Current drinking (%) | 1215 (93.3) | 390 (89.9) | 408 (93.8) | 417 (96.3) |
| Adherence to a healthy diet (%) | 847 (65.3) | 256 (59.1) | 292 (67.4) | 299 (69.4) |
| C-reactive protein, mg/L | 3.41 (5.72) | 4.42 (6.78) | 3.15 (5.08) | 2.68 (4.98) |
| Self-reported health (%) |  |  |  |  |
| Good | 794 (61.2) | 219 (50.7) | 278 (64.2) | 297 (68.8) |
| Fair | 383 (29.5) | 140 (32.4) | 133 (30.7) | 110 (25.5) |
| Poor | 120 (9.3) | 73 (16.9) | 22 (5.1) | 25 (5.8) |
| Waist-to-hip ratio | 0.86 (0.09) | 0.87 (0.09) | 0.86 (0.08) | 0.87 (0.08) |

Abbreviations: MVPA, moderate-to-vigorous physical activity.

**Figure 1.**
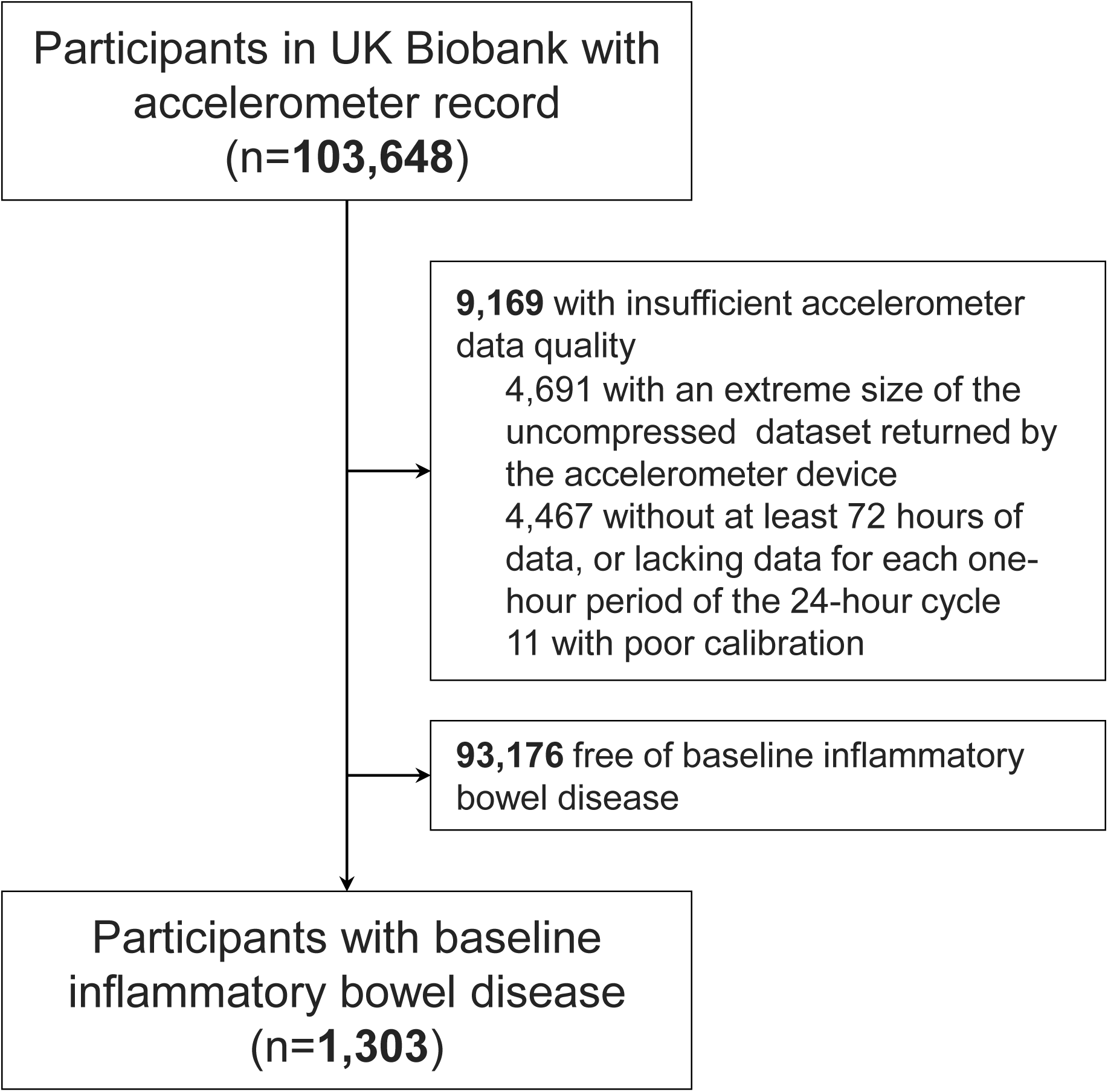
Flow chart of the study.

As shown in **Table 2**, after adjusting for a wide range of sociodemographic and lifestyle factors, we found that higher MVPA duration was associated with reduced bowel resection risk (hazard ratio [HR] per 1-standard deviation [SD], 0.67; 95% confidence interval [CI], 0.47-0.96) and mortality (HR per 1 SD, 0.71; 95% CI, 0.53-0.96). Participants in the highest tertile of MVPA duration had a 56% lower risk of bowel resection surgery (95% CI, 14%-78%) and a 51% lower mortality risk (95% CI, 11%-73%) compared to those in the lowest tertile. No non-linear association between MVPA duration and bowel resection risk was observed (*P* _non-linear_ = 0.13, **Figure 1B**). However, analysis using restricted cubic splines revealed a non-linear relationship between MVPA duration and mortality, characterized by a reverse J-shape and a potentially smaller reduction in risk beyond approximately 58 min/day (*P* _non-linear_ = 0.02, **Figure 1B**). In terms of MVPA pattern, the weekend warrior pattern was associated with a reduced risk of bowel resection (HR, 0.28; 95% CI, 0.12-0.65) but not with reduced mortality (HR, 0.79; 95% CI, 0.44-1.42) compared to the inactive. The regularly active pattern was inversely associated with the risks of bowel resection (HR, 0.37; 95% CI, 0.19-0.69) and mortality (HR, 0.53; 95% CI, 0.31-0.91) (**Table 2**).

**Table 2.**
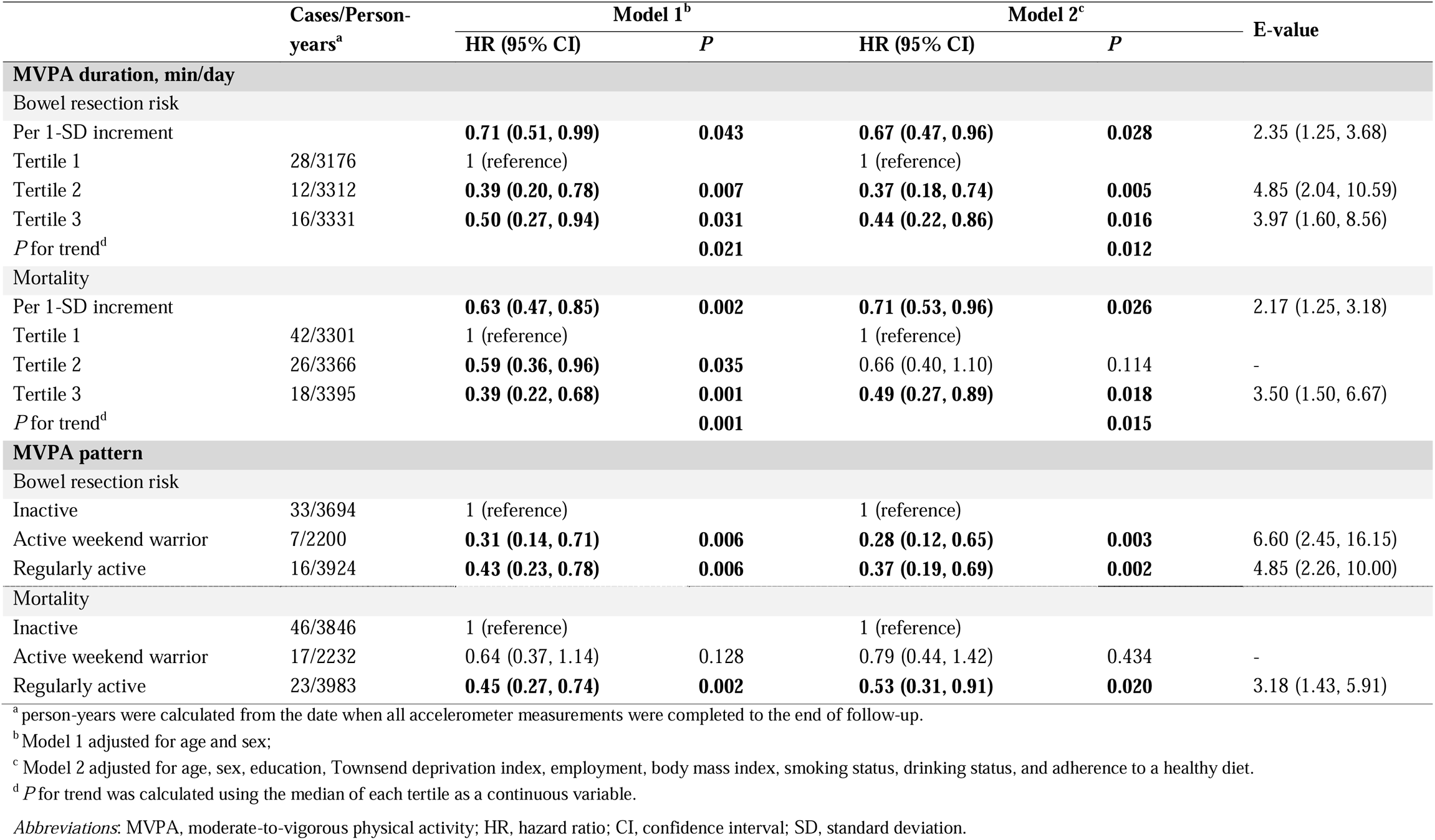
Associations of moderate-to-vigorous physical activity with bowel resection risk and mortality.

In age-stratified analyses, the inverse associations between MVPA duration and mortality were more pronounced in individuals over 60 compared to those under 60 (> 60 years old, HR per 1-SD, 0.47; 95% CI, 0.30-0.74; ≤ 60 years old, HR per 1-SD, 1.04; 95% CI, 0.70-1.55), with both multiplicative and additive interactions (*P* ≤ 0.04) observed. Additionally, in analyses stratified by time since IBD diagnosis, the inverse associations between MVPA duration and bowel resection risk were more pronounced in those with IBD for less than 15 years (> 15 years, HR per 1-SD, 0.87; 95% CI, 0.59-1.28; ≤15 years, HR per 1-SD, 0.34; 95% CI, 0.15-0.77), also showing both multiplicative and additive interactions (*P* ≤ 0.02). In contrast, the association of MVPA patterns with both bowel resection and mortality remained consistent across strata of sex, age, and time since IBD diagnosis (all P > 0.05) (**Figure 3**). Results from separate analyses in individuals with CD and UC were largely consistent with the main findings (**Table S3**).

**Figure 2.**
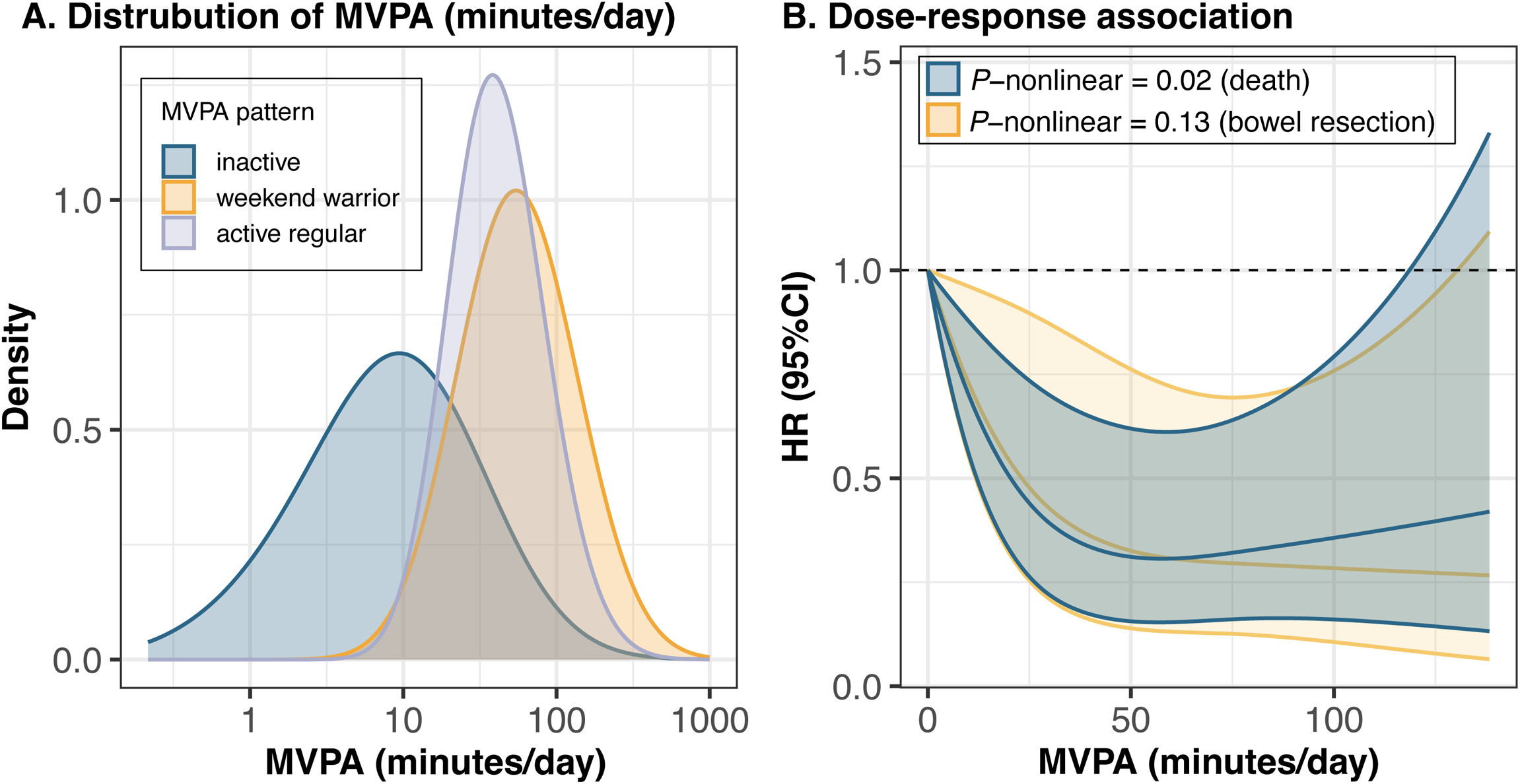
Distribution of daily moderate-to-vigorous physical activity (A) and dose-response associations of daily moderate-to-vigorous physical activity with mortality and bowel resection risk (B). Abbreviations: MVPA, moderate-to-vigorous physical activity; HR, hazard ratios.

**Figure 3.**
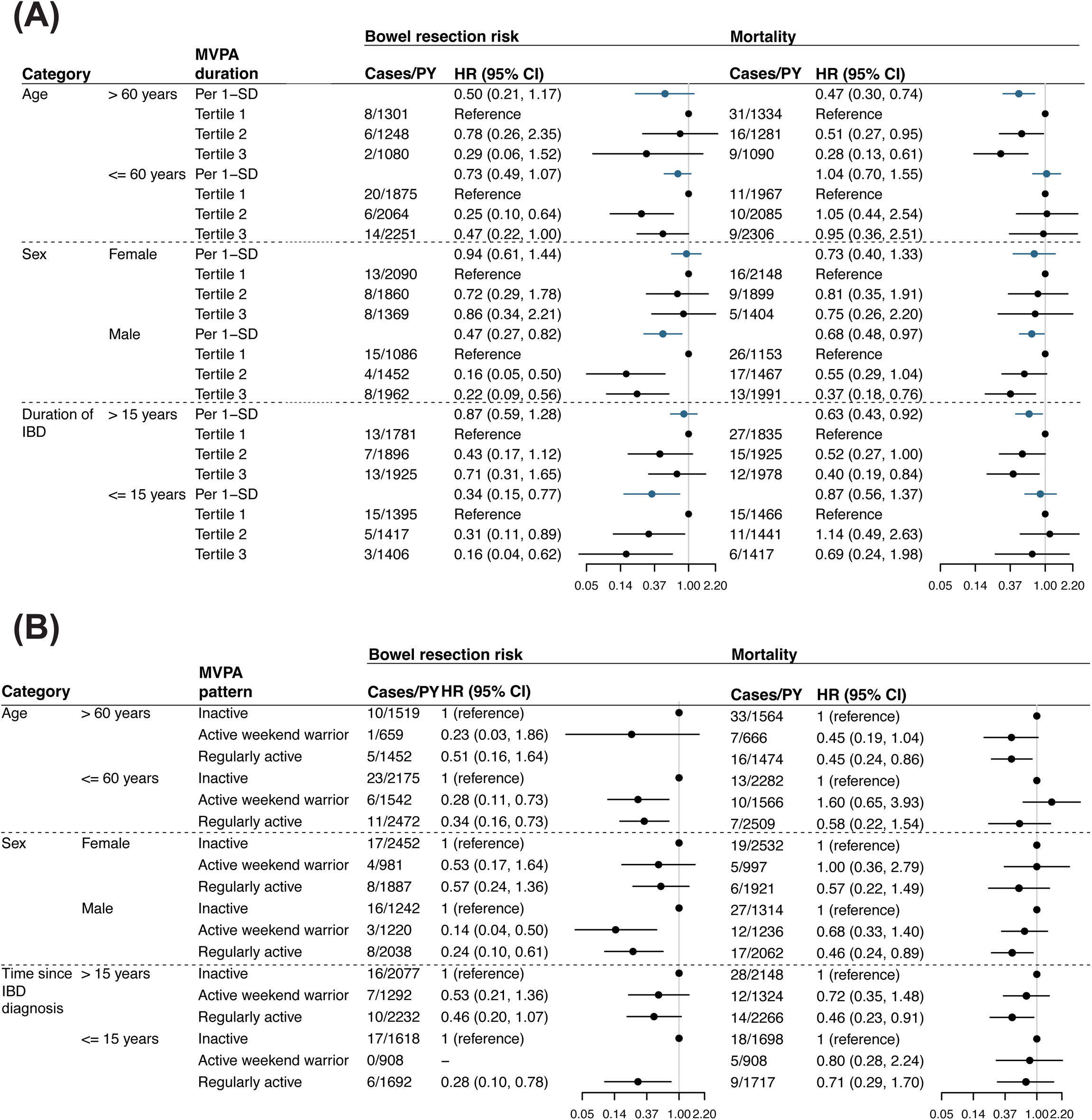
Associations of moderate-to-vigorous physical activity duration (A) and pattern (B) with bowel resection risk and mortality stratified by age, sex, and time since IBD diagnosis. Model adjusted for age, sex, education, Townsend deprivation index, employment, body mass index, smoking status, drinking status, and adherence to a healthy diet. Only two significant interactions were found: between MVPA duration and age on the risk of mortality (p for multiplicative interaction = 0.04, p for additive interaction= 0.002) and between MVPA duration and time since IBD diagnosis on the risk of bowel resection (p for multiplicative interaction = 0.02, p for additive interaction= 0.004). ^a^ person-years were calculated from the date when all accelerometer measurements were completed to the end of follow-up. Abbreviations: MVPA, moderate-to-vigorous physical activity; HR, hazard ratio; CI, confidence interval; SD, standard deviation.

Results were consistent, though a little attenuated, when we repeated analyses excluding participants who had the outcome of interest within one year of follow-up, and in separate analyses additionally adjusted for C-reactive protein, self-reported health, WHR, common IBD-related medications, or prior history of bowel resection surgery (Supplementary **Table S4** and **S5**). ^37^

## Discussion

In this prospective cohort study, we comprehensively investigated the effects of both duration and pattern of accelerometer-measured MVPA on IBD prognosis. Our results indicate that longer durations of MVPA are linearly associated with reduced risks of bowel resection surgery, with per SD increment in duration corresponding to a 33% reduction in bowel resection risk. By contrast, the dose-response relationship between MVPA duration and mortality was nonlinear and appeared to reach a plateau at approximately 58 min/day. Compared with the inactive group, both the active weekend warrior pattern and regularly active pattern were associated with lower bowel resection risk, whereas only the regularly active pattern was associated with reduced mortality.

To our knowledge, no previous study has examined the relationship between objectively measured physical activity and prognosis of IBD in large-scale cohorts. Existing literature on physical activity and IBD prognosis, primarily based on self-reported data, supports the benefits of physical activity. Some small observational studies and randomized controlled trials have reported the potential short-term benefits of physical activity on individuals with IBD, including reduced disease activity,^16,38,39^ improved quality of life,^40^ and improved psychological issues^41^. However, few studies have investigated the role of higher intensities of physical activity, such as MVPA. A study on 242 IBD patients found that both self-reported walking and MVPA were independently associated with physical health-related quality of life, particularly higher volume of MVPA above 150 min/week and walking above 60 min/week.^40^ The result supports the impact of MVPA on individuals with IBD. In addition, there is a lack of evidence on long-term prognostic outcomes such as surgery and mortality. Only one cohort study focused on healthy lifestyle involving 363 CD patients and 465 UC patients found that higher self-reported physical activity, measured in metabolic equivalent task values per week, was linked to lower all-cause mortality (HR for quintile 5 vs. quintile 1, 0.35; 95% CI, 0.19-0.65), which is in agreement with our analysis.^42^

Our study fills these gaps by using accelerometer data to measure physical activity objectively and provides important insights into the effect of MVPA on IBD prognosis. One important finding of the present study is the significant association between MVPA and both reduced mortality and bowel resection risk. To date, national organizations supporting individuals with IBD, such as the Crohn’s Colitis Foundation and Crohn’s and Colitis Canada, advocate for low-to-moderate exercise, including walking, treadmill running, bicycling, and swimming.^43^ However, Our finding challenges the current consensus for individuals with IBD in which they were only encouraged to safely participate in low-to-moderate exercise. According to our findings, the standard guidelines for healthy individuals, recommending at least 150 minutes of MVPA per week, are equally applicable to individuals with IBD. Furthermore, the similar associations of MVPA and bowel resection risk in analyses additionally adjusted for baseline bowel resection history suggest that participating in MVPA seems to offer comparable effectiveness in preventing adverse outcomes, even for those with a history of bowel resection.

Another key finding is the varying impact of different MVPA patterns on health outcomes. Despite similar overall MVPA volumes, only the regularly active pattern is linked to reduced mortality, highlighting that the timing and distribution of physical activity are critical for mortality reduction in individuals with IBD. The differential effects of the two activity patterns on mortality and surgery risk may stem from different driving factors of the outcome. Surgical risk is primarily associated with direct bowel injury and disease activity, whereas mortality is more likely to be closely linked to systemic inflammation and comorbidities, which may be more related to activity patterns. Regular, evenly distributed activity helps maintain lower levels of systemic inflammation more effectively than a concentrated activity pattern, thereby offering better long-term mortality benefits. The present study suggests that a more evenly spread pattern of physical activity is the optimal choice for individuals with IBD. However, less frequent activity sessions, which may be easier to practice, still provide health benefits compared to being inactive.

The strengths of this study include its large sample size, extended follow-up period, and the objective measurement of physical activity through accelerometry. The study also benefited from comprehensive adjustment for important covariates and robust findings across multiple sensitivity analyses, enhancing the reliability of the results. However, there are some limitations. Firstly, physical activity was assessed only once over a 7-day period at baseline, which may not accurately reflect changes in activity patterns over time. Variations in MVPA, driven by factors such as age, health status, or lifestyle changes, could influence the strength of its association with health outcomes. However, prior research has examined the long-term reproducibility of accelerometer-based measurements and found that a 7-day accelerometer assessment of physical activity offers moderate stability over time.^27,28^ Secondly, the study population was predominantly white (>90%), which may limit the generalizability of the findings to other racial or ethnic groups. However, the large size and diverse exposure metrics of the UK Biobank enable valid broadly applicable assessment of exposure-disease relationships.^45^ Thirdly, despite rigorous adjustments, residual confounding by unmeasured factors cannot be completely ruled out, as is the case with all observational studies. We calculated E-values to quantify the potential impact of unmeasured confounding. The large E-values obtained in our analyses suggest that substantial unmeasured confounding would be necessary to nullify the observed associations, indicating robust findings (**Table 2**).^35^

In conclusion, device-measured MVPA duration was associated with lower risks of bowel resection surgery and mortality. An active weekend warrior pattern, characterized by 1 or 2 sessions per week, is inversely associated with bowel resection risk, while a regularly active pattern is inversely associated with both bowel resection risk and mortality. The findings suggest that regular participation in MVPA may be an effective strategy to improve long-term outcomes and mitigate adverse prognoses in individuals with IBD. Recommendations to increase MVPA as a potential management strategy for IBD should be explored.

## Data Availability

Data availability statement: the datasets analysed during the current study are available in a public, open-access repository (https://www.ukbiobank.ac.uk/).

## Additional information

## Acknowledgment

This research was conducted using the UK Biobank study under Application Number 73595. We want to thank all UK Biobank participants and the management team for their participation and assistance. This research was not preregistered in an independent, institutional registry.

## Credit authorship contribution statement

All authors read and approved the final manuscript.

Zixuan He (Conceptualization: Equal; Methodology: Equal; Writing - original draft: Supporting; Writing - review & editing: Equal); Yuhao Sun (Conceptualization: Equal; Methodology: Equal; Formal analysis: Leading; Writing - original draft: Equal; Writing - review & editing: Equal); Hanyi Huang (Conceptualization: Supporting; Visualization: Equal; Writing - original draft: Equal; Writing - review & editing: Supporting); Yilong Liu (Writing - review & editing: Supporting); Lintao Dan (Formal analysis: Supporting; Writing - original draft: Supporting; Writing - review & editing: Supporting); Xixian Ruan (Writing - original draft: Supporting; Writing - review & editing: Supporting); Tian Fu (Writing - original draft: Supporting; Writing - review & editing: Supporting); Zhaoshen Li (Writing - review & editing: Supporting); Xiaoyan Wang (Conceptualization: Equal; Data curation: Equal; Funding acquisition: Equal; Writing - review & editing: Supporting); Jie Chen (Conceptualization: Leading; Data curation: Equal; Writing - review & editing: Equal); Yu Bai (Conceptualization: Equal; Funding acquisition: Leading; Writing - review & editing: Supporting).

## Ethical approval

The manuscript wasn’t previously published or is under consideration for publication elsewhere. The ethical approval was granted for the UK Biobank by the National Health Service National Research Ethics Service (REC reference: 21/NW/0157).

